# Water, sanitation, and hygiene in selected health facilities in Ethiopia: risks for healthcare acquired antibiotic resistant infections

**DOI:** 10.1101/2023.07.12.23292549

**Authors:** Teshome Bekele, Abebe Assefa, Lavuun Verstraete, Adey Feleke Desta, Taha Al-Mulla, Kitka Goyol, Kaleab Baye

## Abstract

**Background:** Inadequate water, sanitation and hygiene (WASH) in health facilities and the low adherence to infection control protocols can increase the risk of hospital-acquired (nosocomial) infections, which in turn can increase morbidity and mortality, health care cost, but also contribute to increased microbial resistance.

**Objectives:** The study aimed to assess WASH facilities and practices, and levels of nosocomial pathogens in surface and water samples collected from selected health facilities in Oromia Region and Southern, Nations and Nationalities and Peoples Region (SNNPR).

**Methods:** WASH in health care facilities in Bulle and Doyogena (SNNPR) and Bidre (Oromia) were assessed through interviews and direct observations (n= 26 facilities). Water and surface samples were collected from major hospitals and health centers. A total of 90 surface swabs and 14 water samples were collected from which a number of bacteria (n=224) were identified, characterized and tested for antimicrobial susceptibility.

**Results:** Water supply, toilet facilities, and waste management procedures were suboptimal. Only 11/26 of the health facilities had access to water at the time of the survey. The lowest hand-hygiene compliance was for Bidre (4%), followed by Doyogena (14%), and Bulle (36%). Over 70% of the identified bacteria were from four categories: *Staphylococcus* spp, *Bacillus* spp, *E. coli,* and *Klebsiella spp.* These bacteria were also found in high risk locations including neonatal intensive care units, delivery and surgical rooms. Antimicrobial susceptibility was detected in ≥ 50% of the isolates for penicillin, cefazolin, ampicillin, oxacillin, and cotrimoxazole, and ≥ 50% of the isolates displayed multi-drug resistance.

**Conclusion:** Investing in WASH infrastructures, promotion of handwashing practices, implementing infection prevention and control (IPC) measures and antibiotic stewardship is critical to ensure quality care in these settings.

## 1. Background

Hospital-acquired infections can increase morbidity and mortality, increase health care cost due to prolonged stay, and also contribute to increased microbial resistance due to the widespread occurrence of multi-drug resistant pathogens in health facilities. Approximately, one-third of neonatal deaths annually (680,000) are caused by infections (UNIGME, 2021). The share of hospital-acquired infection to this remains uncertain, but earlier studies have shown that rates of neonatal infections among hospital-born children in low income countries are 3-20 times higher than those in higher income countries (Zaidi et al., 2005). Most of these infections were present soon after birth and were resistant to antibiotics. Inadequate water, sanitation and hygiene (WASH) and the low adherence to infection control protocols, unsafe waste management, exacerbated by the overcrowding of health facilities increase the risk for hospital-acquired infections (Guo et al., 2017; Schwab et al., 2012).

Recent estimates suggest that hospital-acquired infections affect about 8% of patients in regular wards and more than half of patients admitted in intensive care units in low income settings (Vincent et al., 2009; Alp et al., 2019). A recent study from Jimma University Medical Center reported a prevalence of hospital-acquired infection of 19%, and the risk was significantly higher in those that received surgical procedures(Ali et al., 2018). A study conducted in rural health care facilities in Ethiopia, Kenya, Mozambique, Rwanda, Uganda and Zambia, reported that less than 50% of the surveyed facilities had access to: improved water sources on their premises, improved sanitation, hand washing facilities with constant access to water and soap (Guo et al., 2017). In Ethiopia, only an estimated 30% of health facilities have access to basic water services. However, such data is scarce for lower level health facilities such as *woreda* (district) health centers and health posts, where the problem may be even more significant.

Understanding the magnitude of nosocomial pathogens and their antimicrobial resistance would help design interventions that improve WASH and infection prevention control in health care facilities, but will also contribute to improving the quality of health care delivered. Therefore, the present study aimed to assess WASH facilities and practices, and levels of nosocomial pathogens in hand-touch sites in selected health facilities in Oromia and SNNPR.

## 2. Methods

### Study area and design

This study is reporting on a baseline assessment conducted in health facilities in Bidre town in Bale zone in Oromia Region, Bulle town in Gedeo zone in SNNPR region and Doyogena in Kembata-Tembaro zone in SNNPR region. The WASH assessments included all health facilities that were functional at the time of the survey (i.e. health post, health centers, and hospitals). Surface and water sample were primarily collected from hospitals and health centers.

### Assessment of WASH in health facilities

A WASH observational checklist was developed based on international standards— WHO/UNICEF core questions for water and sanitation, IPC and WASH common indicators (WHO, 2019). All questionnaires and checklists were translated into Amharic/Oromifaa and were pretested prior to the interviews. The checklist allowed the collection of information on the prevailing sanitary conditions, access to water and hand-washing facility, as well as hand-washing and waste disposal practices. The WHO protocol on monitoring fulfilment of opportunities for hand-hygiene was used to assess the health personnel’s adherence to hand-hygiene guidelines (WHO, 2009).

### Surface and water sample collection

Sample collection was preformed following the United States Center for Disease Control and Prevention (CDC) and Public health England guidelines (CDC, 2017; Willis et al., 2013). Surface sample collection was performed using sterile cotton swabs. The swabs were first moist in sterile normal saline solution. The samples were collected from surfaces including beds, door handles, walls, gowns, autoclaves, tables, and chairs. The sampling areas included out-patients departments, different wards, pharmacy, laboratories, receptions, toilets and cafeterias in the health facilities.

Water samples were collected from sources from which the health facilities obtain water for washing, drinking and other activities in the healthcare settings. A total of 15 hospital water samples were collected from delivery wards, medical ward, tanker, and bore-hole and rainwater collection systems. Overall, 59 water samples were collected from all health facilities, including health centers and health posts.

### Sample handling and transportation

The collected surface samples were immediately put in Amies transport media and kept in pre-cooled ice box and transported to SNNP region’s Public Health Institute laboratory. On arrival at the laboratory, the surface samples were transferred to the nutrient broth and enriched overnight at 37 LC. After an overnight incubation, the samples were inoculated on blood agar and MacConkey agar plates and put overnight at 37LC. In case of no growth after an overnight culture, the plates were incubated for an additional 24 hours.

The water samples were assessed for their safety using modified Method 9215 to enumerate heterotrophic bacteria and membrane filtration technique for Gram-negative bacteria (APHA, 2005). To enumerate heterotrophic bacteria, one ml of each water sample was pipetted into a sterile petri dish. After thoroughly mixing, the melted MacConkey agar was poured into the dish. The melted medium was mixed thoroughly with the sample and solidified. The plates were incubated for 48 hours at 37°C.

The Gram-negative bacteria were counted by filtrating 100 ml water samples through 0.45 µm pore size-47 mm, and cellulose nitrate membranes using the modified ISO 9308-1 protocol (ISO, 2014). The samples were incubated on MacConkey agar for 24 hours at 37°C. All results of Gram-negative bacteria were expressed as colony forming units per 100 mL water. The bacterial colonies were collected and put in Trypticase Soy Broth containing 20 % glycerol and were transported to the National bacteriology and mycology Reference Laboratory (NRL) at the Ethiopian Public Health Institute, where they were stored in deep-freeze until further analyses.

### Bacterial isolation and identification

The bacteria were refreshed by culturing on three different culture media: i) 5% sheep blood agar plate, ii) MacConkey agar plate, and iii) Mannitol salt agar plate. Colony appearance on culture plates, microscopic examination, and biochemical tests were used to identify Gram-positive and Gram-negative bacteria.

#### Identification of Gram-positive cocci

The common Gram-positive cocci are *Staphylococcus spp* and *Streptococcus spp*. We used Blood agar and Mannitol salt agar media for isolation of *Staphylococcus spp*: The culture plates were incubated in air at 37°C for 24 hour. Colony morphology on culture plates and microscopic examination for Gram-positive cocci in clusters were used for initial *Staphylococcus spp* identification. Catalase and coagulase tests were used to classify *Staphylococcus spp* into *Staphylococcus aureus* and coagulase negative *Staphylococcus*. All *Staphylococci* are Catalase positive and only *S. aureus* is coagulase positive.

*Streptococcus* spp were identified based on colony morphology on: i) blood agar plates (beta hemolytic, alpha hemolytic and non-hemolytic), ii) microscopic examination for Gram-positive in chin, and iii) different biochemical tests. Negative catalase test differentiated *Streptococcus* spp from *Staphylococci Bacillus* spp.

Blood agar with 5 % sheep blood media was used for the bacteria isolation. Colony morphology on the culture plates and gram stain were used for the bacterial identification. To differentiate *Bacillus cereus* from other *Bacillus* species we used citrate test which is only positive for *B. cereus*.

#### Identification of Gram-negative bacilli

The common gram negative bacteria are generally divided into two major categories: Fermenters and non-fermenters. Fermenters gram-negative bacilli utilize lactose and become pink color colonies on MacConkey agar while non-fermenters cannot utilize lactose and they are colorless colonies on MacConkey agar plate. Biochemical tests such as Triple Sugar Iron Agar (TSI), urea, citrate, Sulfide Indole Motility (SIM) medium, growth in Lysine Iron Agar (LIA), and oxidase were additionally used to identify Gram-negative bacteria.

### 2.4 Antimicrobial susceptibility testing

The antimicrobial Susceptibility Tests (AST) were performed based on the Kirby–Bauer disk diffusion method on Mueller-Hinton agar (MHA) as recommended by clinical and laboratory standard Institute (CLSI) for all Gram-negative bacteria and *Staphylococcus* species (CLSI, 2017). Well-isolated three to four colonies were emulsified in a tube containing sterile normal saline and the turbidity adjusted to 0.5 McFarland standards. The emulsified bacterial suspension was uniformly streaked on MHA plates using sterile cotton swabs, on which the antibiotic disks were applied and incubated for 18-24 hours at 37 LC. The antibiotic agents tested in this study were ampicillin (10 µg), amoxicillin-clavulanic acid (20/10 µg), pepracillin/ tazobactum, cefazolin (30 µg), cefuroxime (30 µg), cefotaxime (30 µg), ceftazidime (30 µg), cefepime (30 µg), cefoxitin (30 µg), ciprofloxacin (5 µg), amikacin (30 µg), meropenem (10 µg), chloramphenicol, tetracycline, cotrimoxaxole, and penicillin. Penicillin and cefoxitin were tested only for *Staphylococcus* species and the result of oxacillin was determined from cefoxitin breakpoint. Antibiotic susceptibility results were interpreted according to the CLSI zone size interpretive standards (CLSI, 2017). Intermediate results were considered to be resistant.

Multidrug resistance (MDR) was defined according to guidelines compiled by the European Center for Disease prevention and Control (ECDC) and the Centers for Disease Control and Prevention (CDC) (Magiorakos et al., 2012). Accordingly, bacterial isolates that were resistant to at least one agent in three different antimicrobial categories were considered as MDR.

### 2.5 Quality Assurance

All media, biochemical reagents, gram stain reagents and antibiotic disks were checked for their quality using standards ATCC strains. Standard ATCC quality strains used for this study were *S. aureus* ATCC^®^ 25923, *E. coli* ATCC^®^ 2592, P. aeruginosa ATCC 27853.

### 2.6 Data analysis

The data were entered and cleaned using Microsoft Excel and were exported to SPSS version 20.0 for further analyses. The frequencies of bacterial isolates and antimicrobial susceptibility were calculated. Mean and frequencies (percentage) were used to present descriptive data.

### 2.7 Ethics

Ethical clearance was obtained from the Institutional Review Board of the College of Natural and Computational Sciences of Addis Ababa University. Informed consent was obtained prior collection of data.

## 3. Results

WASH assessments were conducted in 26 health facilities in Bulle and Doyogena (SNNP Region) and in Bidre (Oromia Region). The assessments included hospitals (n=3), health posts (n=13), clinics (n= 8), and health centers (n=2; **Table 1**). A great majority of the health facilities relied on tanker trucks for their water supply (**Table 2**). At the time of the survey, piped water supply was available in only 11 of the 26 health facilities. Open pit latrines (14/26) were the commonest type of toilet and only in 8 out of the 26 facilities, the toilets were accessible for people with limited mobility. Infectious waste was primarily dumped into an open/protected pit, incinerated, and added to other wastes. Sharp waste was mostly collected for off-site disposal, autoclaved, or incinerated. Only 10 of the 26 assessed health facilities had guidelines on standard precautions for infection prevention control (IPC). Only six had cleaning protocols available, and only in one health facility, the staff responsible for cleaning received training. Environmental disinfectant was only available in only 8 of the 26 health facilities.

**Table 1.**
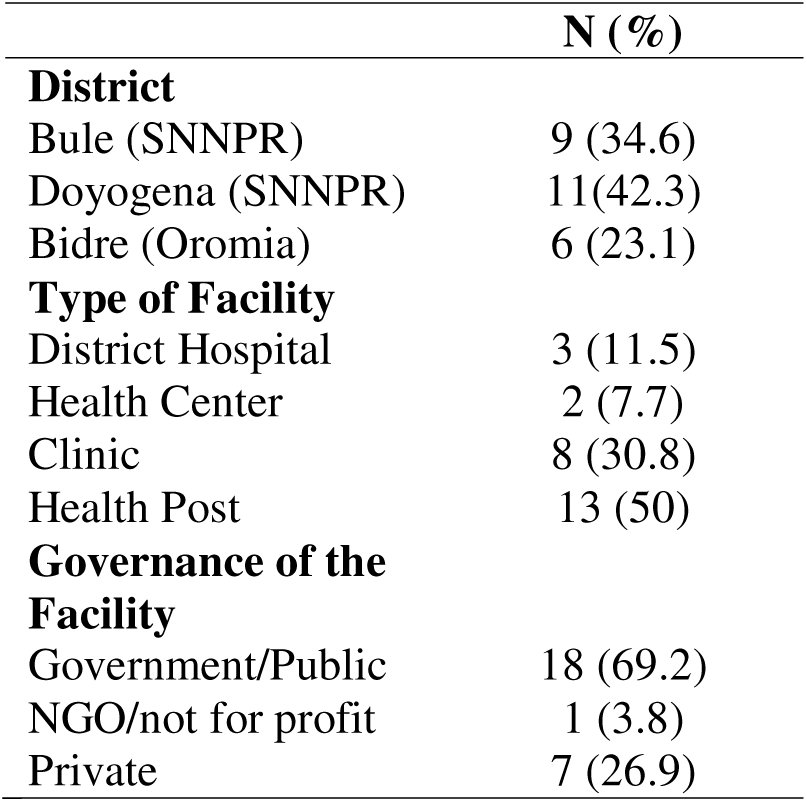
Characteristics of the health facilities.

**Table 2.**
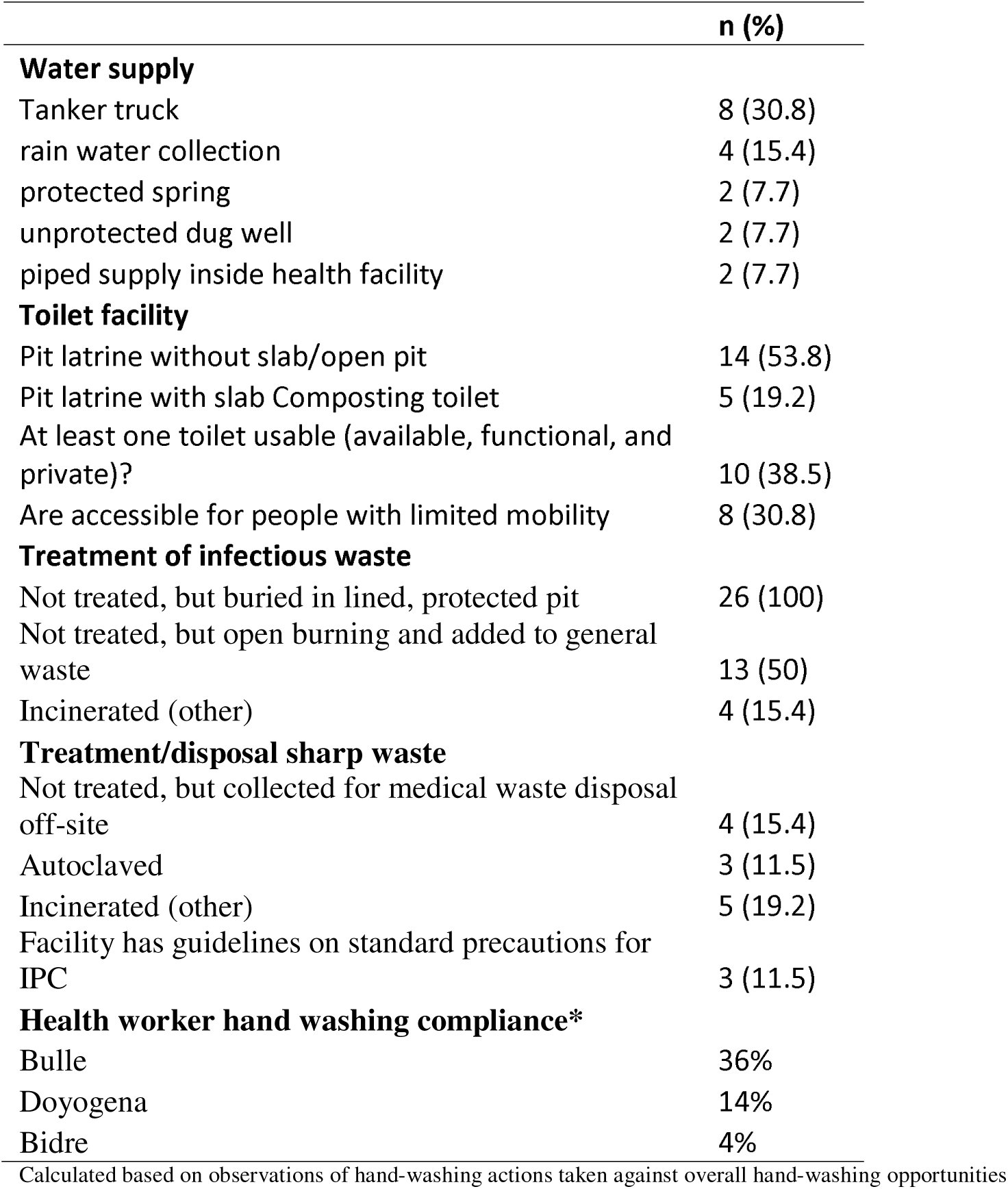
Water, Sanitation, and Hygiene of the assessed health facilities.

Hand-hygiene opportunities were directly observed (1194 ± 326 min) and evaluated using the WHO checklist to assess compliance (**Table 2**). Hand-hygiene opportunities were: i) before touching a patient; ii) before a procedure; iii) after body fluid exposure/risk; iv) after touching a patient; v) after touching a patient’s surrounding. Hand-hygiene compliance was overall low, but varied by site. The lowest compliance was for Bidre (4%), followed by Doyogena (14%), and Bulle (36%).

A total of 90 surface swabs and 14 water samples were collected from which a number of bacteria (n=224) were identified (**Table 3**). Over 70% of the identified bacteria were from four categories: *Staphylococcus* spp, *Bacillus* spp, *E. coli,* and *Klebsiella spp.* These bacteria were the most widely distributed and were also found in high risk locations including neonatal intensive care units, delivery and surgical rooms (**Table 4**). More details on the identified bacteria by study sites, location and sample source can be found in the **supplemental table S1-S3 and figure S1**. **Figure 1** presents the antimicrobial resistance of the identified bacterial isolates. Antimicrobial susceptibility was detected in 50% or more of the isolates for penicillin, cefazolin, ampicillin, oxacillin, and cotrimoxazole. More than 50% of the isolates displayed multi-drug resistance, defined as resistant to at least one agent in three different antimicrobial categories.

**Figure 1.**
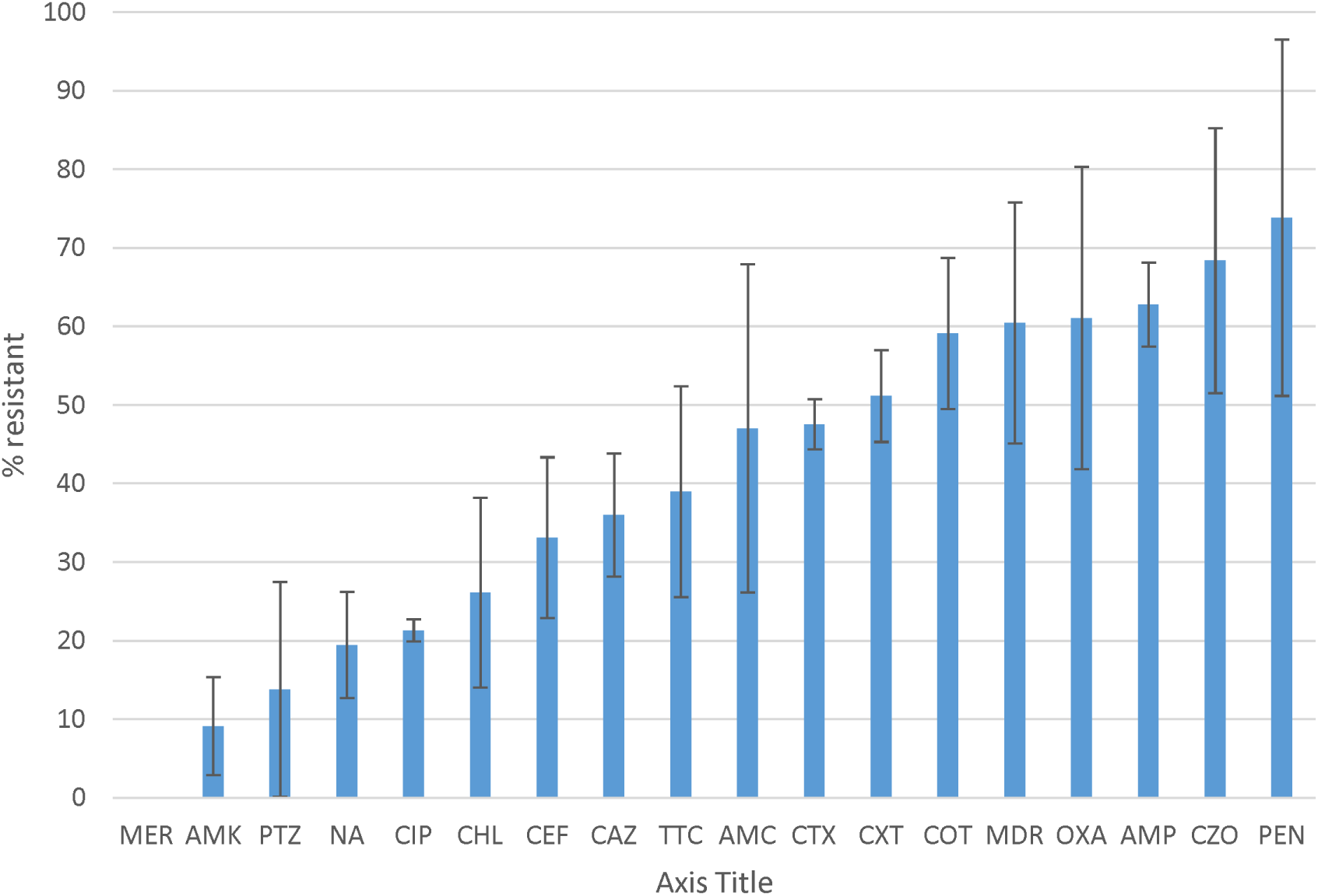
Antibiotic susceptibility profile and multidrug resistance of the identified bacteria. Percentage of resistance of bacterial isolates identified from healthcare surface environmental and water samples according to the CLSI disk diffusion breakpoints. Resistance was defined as isolates with intermediate resistance and complete resistance inhibition zone size. Antibiotics tested were ampicillin (AMP), amoxicillin-clavulanate (AMC), pepracillin with tazobactum (PTZ), cefazolin (CZO), cefuroxime (CXT), ceftazidime (CAZ), cefepime (CEF), cefotaxime (CTX), ciprofloxacin (CIP), nalidixic acid(NA), chloramphenicol (CHL), Cotrimoxazole (COT) Amikacin (AMK), Meropenem (MER),tetracycline (TTC), Penicillin (PEN) and oxacillin (OXA). MDR is to indicate the rate of Multidrug resistant bacterial isolates.

**Table 3.** Identification of bacteria from surface swabs and water samples.

| Bacteria Isolates | Bulle | Doyogena | Bidre | Total |
| --- | --- | --- | --- | --- |
|  | n (%) |  |  | N (%) |
| *E. hermani* | 1(1.4) | - | - | 1 (0.4) |
| *Providencia* spp | - | 1 (1.3) | - | 1 (0.4) |
| *Proteus mirabilis* | 1(1.4) | - | - | 1 (0.4) |
| *Enterococcus* spp | - | - | 2(2.5) | 2 (0.9) |
| *Morganella* spp | - | 2 (2.7) | - | 2 (0.9) |
| *Pseudomonas* spp | - | 6 (8.0) | 1(1.3) | 7 (3.1) |
| *Alcaligenes* spp | - | 8 (10.7) | - | 8 (3.5) |
| *Citrobacter* spp | 2(2.9) | 5 (6.7) | 2(2.5) | 9 (4.0) |
| *Enterobacter* spp | 1(1.4) | 3 (4.0) | 6(7.6) | 10 (4.5) |
| *Acinitobacter* spp | 7 (10.0) | 5 (6.7) | 6 (7.6) | 24 (10.7) |
| Rare Non- fermenters | 7 (10.0) | - | 17 (21.5) | 24 (10.7) |
| *Klebsiella* spp | 12 (17.1) | 9 (12.0) | 9(11.6) | 30 (13.4) |
| *E. coli* | 12 (17.1) | 15 (20.0) | 8 (10.1) | 35 (15.6) |
| *Bacillus* spp | 13(18.6) | 14 (18.7) | 9(11.6) | 36 (16.1) |
| *Staphylococcus* spp | 12 (17.1) | 7 (9.3) | 19 (24.1) | 38 (17.0) |
| **Total** | **70 (100)** | **75 (100)** | **79 (100)** | **224 (100)** |

**Table 4.** Identified bacteria by sample source/location.

|  | *Acinetobacter* | *Alcaligenes* | *Bacillus* | *Citrobacter* | *E. coli* | *Edwardians* | *Enterobacter* | *Klebsiella* | *Non-lactose fermenter* | *Pseudomonas* | *Staphylococcus* | *Morganella* | *Providencia* | *S. aureus* | N of different pathogenic bacteria found | N of samples |
| --- | --- | --- | --- | --- | --- | --- | --- | --- | --- | --- | --- | --- | --- | --- | --- | --- |
| Adult OPD |  |  |  |  |  |  |  | x | x |  | x | x |  |  | 4 | 4 |
| ANC (Health center) |  | x |  |  |  |  |  |  |  |  |  |  |  |  | 1 | 3 |
| Autoclave |  |  |  |  |  |  |  |  |  |  | x |  |  |  | 1 | 1 |
| Card room |  | x | x |  | x |  |  |  |  |  |  |  |  |  | 3 | 1 |
| Delivery room | x | x | x | x |  |  |  | x |  |  | x |  |  | x | 7 | 5 |
| Door handle | x | x | x | x | x |  |  |  |  |  | x |  |  |  | 6 | 5 |
| Emergency | x | x | x | x | x |  | x |  |  | x | x |  | x | x | 10 | 6 |
| Family planning |  | x | x |  |  |  |  |  |  | x |  |  |  |  | 3 | 2 |
| Female medical ward |  | x | x |  | x |  | x |  |  |  | x |  |  |  | 5 | 2 |
| Female surgical |  |  |  |  |  |  | x | x |  |  | x |  |  |  | 3 | 1 |
| Gynecology |  |  |  | x | x |  |  |  |  |  | x |  |  |  | 3 | 2 |
| HC Adult OPD |  |  |  |  |  |  |  |  | x |  | x |  |  |  | 2 | 2 |
| HC ART clinic |  |  |  |  |  |  |  |  | x |  |  |  |  |  | 1 | 1 |
| HC Emergency OPD |  |  |  |  |  |  |  |  | x |  |  |  |  |  | 1 | 2 |
| HC EPI |  |  |  |  |  |  |  |  | x |  | x |  |  |  | 2 | 2 |
| HC laboratory | x |  | x |  | x |  |  |  |  |  | x |  |  | x | 5 | 2 |
| HC water |  |  |  |  |  |  |  |  | x |  |  |  |  |  | 1 | 1 |
| Hospital water |  |  |  |  |  |  |  |  |  |  |  |  |  |  | 0 | 1 |
| Laboratory |  |  |  |  |  |  |  |  |  | x |  |  |  |  | 1 | 3 |
| Male surgical ward |  |  | x | x | x |  |  | x | x | x | x |  |  |  | 7 | 3 |
| Medical ward |  |  | x |  | x |  |  |  |  |  |  |  | x |  | 3 | 1 |
| Neonatal ICU |  |  | x |  |  |  |  |  |  |  |  |  |  | x | 2 | 2 |
| Observation room | x | x | x |  |  |  | x | x |  |  |  |  |  |  | 5 | 2 |
| Obstetric room |  |  |  |  |  |  | x | x |  |  | x |  |  |  | 3 | 1 |
| Operation room |  |  | x |  | x |  | x |  |  |  | x |  |  |  | 4 | 1 |
| Pediatric isolation room |  |  | x |  |  |  | x |  | x |  | x |  |  |  | 4 | 2 |
| Pediatric OPD |  |  | x |  |  |  |  |  | x |  | x |  |  |  | 3 | 5 |
| Pediatric ward |  |  |  |  | x |  | x | x | x | x |  |  |  | x | 6 | 4 |
| Pharmacy |  |  | x |  | x |  |  |  | x |  |  |  |  |  | 3 | 3 |
| Post-natal |  |  | x |  | x |  | x |  |  |  | x |  |  | x | 5 | 3 |
| Pre-term |  |  |  |  | x | x |  |  |  |  |  |  |  |  | 2 | 1 |
| Procedure room | x |  |  |  | x |  |  |  |  |  |  |  | x |  | 3 | 3 |
| Psychiatric room |  |  |  |  |  |  |  |  |  |  | x |  |  |  | 1 | 1 |
| Rain water |  |  |  |  |  |  |  |  |  | x |  |  |  |  | 1 | 1 |
| Reception |  |  | x |  |  |  |  |  | x |  | x |  |  |  | 3 | 4 |
| Stabilization phase I |  |  | x |  |  |  |  | x | x |  | x |  |  | x | 5 | 3 |
| Staff cafeteria |  |  |  |  |  |  | x |  |  |  |  |  |  |  | 1 | 1 |
| Sterilization room |  |  |  |  |  |  |  |  |  |  |  |  |  | x | 1 | 1 |
| Surgical ward |  |  | x |  | x |  |  |  |  |  |  |  |  | x | 3 | 2 |
| Table HP |  | x | x | x | x |  | x |  |  |  |  |  |  |  | 5 | 2 |
| Tanker 1 water |  |  |  | x | x |  |  |  |  | x |  |  |  |  | 3 | 1 |
| Toilet door handle |  |  |  |  | x |  |  |  |  |  |  |  |  |  | 1 | 2 |
| Under 5 OPD HC |  |  |  |  |  |  |  |  |  |  |  | x |  |  | 1 | 6 |
| Under five emergency |  |  | x |  |  |  |  |  |  |  |  |  |  |  | 1 | 1 |
| Wards door |  |  |  |  |  |  |  | x | x |  | x |  |  |  | 3 | 1 |
| Will chair |  |  |  |  | x |  |  |  |  |  |  |  |  |  | 1 | 1 |

ART, antiretroviral therapy; EPI, expanded programme on immunization; HC, health center; ICU, intensive care unit; OPD, outpatient department

## 4. Discussion

Water supply, availability of clean and accessible toilets, as well as infection prevention measures were found suboptimal. Hand hygiene practice by health workers was very low. Consequently, surface swabs and water samples revealed high bacterial contamination, with some of the identified bacteria known for their pathogenicity. These bacteria were also found in highly sensitive areas like surgical rooms, delivery rooms, and neonatal intensive care units (ICUs).

Our findings highlight the need to invest in safely managed water supply, provision of safely managed sanitation services, but also strict hygiene and environmental cleaning in health facilities. Earlier studies assessing 1,318 health facilities in multiple African countries including Ethiopia showed that less than 50% of the facilities had access to improved water sources on premises, improved sanitation, and consistent access to water and soap for handwashing (Guo et al. 2017). A recent meta-analyses of studies on health workers’ handwashing practice in Ethiopia also estimated that 57.87% (95% CI: 44.14–71.61) practiced hand-washing (Gedamu et al., 2021), a figure that is higher than estimates from the current study. This difference may be explained by the rather rigorous evaluation of hand-hygiene practice in this study assessed using the more systematic WHO’s protocol of hand-hygiene opportunities. It can as well suggest that the selected sites have more significant WASH constraints, further justifying their selection for WASH and IPC improvements by the planned intervention.

The poor WASH and IPC conditions observed in the health facilities can greatly impact the quality of the health care provided. First, satisfaction with WASH and IPC conditions can be associated with lower job satisfaction as reported from a recent multi-country study (Fejfar et al., 2021). Second, health facilities with suboptimal WASH and IPC procedures increase the risk for nosocomial infections. Indeed, a recent meta-analyses pooling results from 18 studies in Ethiopia (Alemu et al., 2020), estimated the prevalence of nosocomial infections to be as high as 17% (95% CI 14.10–19.82). This prevalence can be even higher when considering vulnerable sub-groups like neonates, infants and young children. Indeed, studies have shown that hospital-acquired infections contribute significantly to neonatal infections and mortality in low income countries like Ethiopia (Zaidi et al., 2005).

A number of pathogenic bacteria associated with nosocomial infections have been identified from highly sensitive locations like surgical rooms, delivery room, and neonatal ICU. *Klebsiella spp, E. coli, Acinetobacter spp, bacillus spp* and *Staphylococcus spp* were identified in high number of samples collected from various locations. Poor hand-hygiene and bacterial contamination with antimicrobial resistance was a common features of health facilities in all the three sites. More concerning is that a large number of the identified bacteria displayed antibiotic resistance and these same species were reported to be the major pathogens identified in bloodstream isolates (n=11,471) of hospital-acquired neonatal infections (Zaidi et al., 2005). A recent global study showed that most of the bacterial isolates identified in our study were responsible for high rates of deaths associated with antimicrobial resistance, particularly in sub-Saharan African countries (Murray et al., 2022).

The present study has a number of limitations that need to be considered when interpreting our findings. First, this is a cross-sectional study and thus only provides a snapshot of the situation at the time of the survey. Second, the survey happened during the COVID-19 pandemic that in principle would have increased awareness on hand-hygiene because of the nation-wide campaigns. Third, this is a baseline assessment of health facilities selected for WASH/IPC intervention and thus may not be representative. However, evidence from our WASH data is in line with previous assessments and thus can be indicative of situations in similar settings in Ethiopia.

## 5. Conclusion

The health facilities assessed were confronted with serious problems related to WASH. Compliance to hand-hygiene practice by the health care workers was very low. Analyses of environmental and water samples revealed high levels of bacterial contamination. Most of the identified bacteria displayed antimicrobial resistance. Beyond increasing access to health coverage, emphasis should be put to improving infrastructure and services. This requires safely managed water supply, provision of safely managed sanitation services, but also strict hygiene and environmental cleaning in health facilities. Ensuring the supply chain of critical consumables such as soap, chlorine and decontaminants or disinfectants is key, but this will also need to be accompanied by behavioral change on hand hygiene and environmental cleaning practices. A critical element of strengthening health systems should also focus on antibiotic stewardship.

## Acknowledgments

The facilitation offered by the health bureau and the Hospitals of Bidre, Doyo Gena and Bulle is duly acknowledged.

## Funding

Funding for this study was received from UNICE Ethiopia

## Competing interests

None

## Data availability statement

additional data related to this study can be made available upon reasonable request

## Notes

### Competing Interest Statement

The authors have declared no competing interest.

### Funding Statement

UNICEF Ethiopia

### Author Declarations

Ethics committee of Addis Ababa University gave ethical approval for this work.

