## Supplemental for "Water, sanitation, and hygiene in selected health facilities in Ethiopia: risks for healthcare acquired antibiotic resistant infections"

**Supplemental file**

**Table S1** List of bacteria isolated from environmental and water samples from health facilities in Bulle district

| **No** | **Site in the healthcare setting** | **Gram Negative** | **Gram positive** | |
| --- | --- | --- | --- | --- |
| **Surface samples** | | | | |
| 1 | Pediatric OPD (Bed1) | *Acinitobacter* Spp, *Stentophomonas maltophilia* | | *Baccillus* spp |
| 2 | Pediatric OPD (Bed2) | *E. coli*, | |  |
| 3 | Pediatric OPD (table) | *E. coli*, *E. hermani*, *Acinitobacter* spp | |  |
| 4 | Pediatric OPD (table) | *Burkholderia* spp, *Comamonas testosteroni*, Non-fermenter | |  |
| 5 | Chronic OPD | *E. coli*, *Citrobacter* spp | |  |
| 6 | ANC | Non-fermenter, | | *Baccillus* Spp |
| 7 | EPI | *Klebsiella* spp, *E. coli* | |  |
| 8 | Family plan | *K. pneumoniae*, *K. ozeanea* | |  |
| 9 | Delivery | *E. coli*, *Acinitobacter* spp | |  |
| 10 | Reception | *E. coli,* *K. aerogens* | |  |
| 11 | Emergency | *Acinitobacter* Spp | | *S. aureus* |
| 12 | Procedure room | *Pseudomonas* spp | | *S. aureus*, *Bacillus* species |
| 13 | Emergency Trial | *E. cloacae* , *Klebsiella* spp | | *S. aureus*, |
| 14 | NICU | *Klebsiella* spp, | | S. aureus |
| 15 | Delivery room |  | | *Bacillus* spp |
| 16 | Delivery room Gaun | *E. coli* | | *S. aureus* |
| 17 | Postnatal 1 | *E. coli* | | *Bacillus* spp |
| 18 | Postnatal 2 | *E. coli* | | *S. aureus* |
| 19 | Operation Room |  | | *Bacillus* spp |
| 20 | Toilet door hand | *E. coli, Proteus mirablis* | |  |
| 21 | Stabilization 1 | *Klebsiella spp*, Non Lactose fermenter | | *S. aureus* |
| 22 | Stabilization 2 | *E. coli.* | | *S. aureus* |
| 23 | Adult female medical ward | *E. coli*, *Klebsiella spp* | |  |
| 24 | Pediatric medical ward | *Klebsiella spp, Acinitobacter* spp | | *S. aureus* |
| 25 | Surgical ward | *Acinitobacter* spp | | *S. aureus* |
| 26 | Laboratory | *Acinitobacter* spp*,*  *Klebsiella spp* | | *S. aureus, Bacillus* species |
| 27 | Sterilization |  | | *S. aureus*, *Bacillus* spp |
| 28 | Door hand (HP) |  | | *Bacillus* spp |
| 29 | Table (HP) |  | | *Bacillus* spp |
| **Water samples** | | | |  |
| 31  32  33  34  35  36  37 | Delivery ward water  Tanker water  Small tanker water  Medical ward water  Rain water health post  Tanker Seko health post  Borehole Seko health post | *E. coli*  *Non Lactose fermenter, Klebsiella spp*  *Klebsiella spp, Citrobacter spp*  Non Lactose fermenter, | | *Bacillus spp*  *Bacillus spp*  *Bacillus spp* |

ART, antiretroviral therapy; EPI, expanded programme on immunization; HC, health center; ICU, intensive care unit; OPD, outpatient department

**Table S2** Bacteria isolated from environmental and water samples from health facilities in Doyo Gena district

| **No** | **Site in the healthcare setting** | **Gram Negative** | **Gram positive** | |
| --- | --- | --- | --- | --- |
| 1 | Reception | *Acinitobacter* spp | | *Bacillus* spp |
| 2 | Emergency | *Acinitobacter* spp, *Alcaligenes* spp | | *Bacillus* spp, *Staphylococcus* spp |
| 3 | Emergency procedure | *Enterobacter cloacae* | | *Bacillus* spp, *Staphylococcus* spp |
| 4 | Under 5 Emergency | *Acinitobacter* spp | | *Bacillus* spp |
| 5 | NICU | *Acinitobacter* spp | | *Bacillus* spp |
| 6 | medical ward | *E. coli*, *Providencia rettigi* | | *Bacillus* spp |
| 7 | surgical ward | *E. coli, Acinitobacter* spp | |  |
| 8 | female medical ward | *E. coli, K. oxytoca, Alcaligeues faecalis* | |  |
| 9 | Door medical ward | *Citrobacter* spp, *Alcaligeues faecalis* | | *Staphylococcus* spp |
| 10 | Gynecology ward | *E. coli*, *K. aerogenes* | |  |
| 11 | male surgical ward | *E. coli*, *Pseudomonas aeruginosa* | | *Staphylococcus* spp |
| 12 | pediatric ward | *E. coli*, *Pseudomonas aeruginosa* | |  |
| 13 | Laboratory | *E. coli*, *Pseudomonas aeruginosa* | | *Baccillus* Spp |
| 14 | Card room | *E. coli* | | *Baccillus* Spp |
| 15 | Pharmacy | *E. coli* | | *Baccillus* Spp |
| 16 | family planning | *Pseudomonas aeruginosa, Alcaligenes* spp | | *Baccillus* Spp |
| 17 | ANC | *Klebsiella aerogenes* | |  |
| 18 | Operation Room | *E. coli, Enterobacter cloacae* | | *Staphylococcus* spp |
| 19 | Delivery | Citrobacter spp, Klebsiella spp | | *Staphylococcus* spp |
| 20 | Door Zeraro HP | *K. oxytoca, Alcaligenes* spp | |  |
| 21 | Table Zeraro HP | *E. coli*, *Citrobacter* spp | |  |
| 22 | Door Leino HP | *E. coli* | | *Baccillus* Spp |
| 23 | Table Leino HP | *Enterobacter s*pp, *Alcaligenes* spp | | *Baccillus* Spp |
| 24 | Delivery Sararo HC | *K. oxytoca* | | *Staphylococcus* spp |
| 25 | Emergency SararoHC | *K. pneumonia, Citrobacter spp* | |  |
| 26 | Abortion care SararoHC | *K.pneumoniae* | |  |
| 27 | ANC and FP Sararo HC | *Alcaligenes facalis* | |  |
| 28 | Reception sararo HC |  | |  |
| 29 | Under 5 OPD Sararo HC | *Morganella* spp | | *Baccillus* Spp |
| 30 | Adult OPD Sararo HC | *Morganella* spp | | *Baccillus* Spp |
| **Water samples** | | | | |
| 31 | Hospital water tanker 1 | *E. coli*, *Citrobacter* spp | |  |
| 32 | Hospital water tanker 2 | *K. pneumoniae, Pseudomonas* spp | |  |
| 33 | HC water (rain water) | *E. coli*, *Pseudomonas* spp | |  |

ART, antiretroviral therapy; EPI, expanded programme on immunization; HC, health center; ICU, intensive care unit; OPD, outpatient department

**Table S3** List of bacteria isolated from environmental and water samples from health facilities in Bidre district

| **No** | **Site in the healthcare setting** | **Gram Negative** | **Gram Positive** |
| --- | --- | --- | --- |
| 1 | Emergency | *Acinitobacter* spp, *Pseudomonas* spp, | *Bacillus* spp |
| 2 | Observation | *Enterobacter cloacae*, *Klebsiella* spp | *Bacillus* spp |
| 3 | Procedure | *E. coli, Acinitobacter* spp*,* |  |
| 4 | Wards Door | *Acinitobacter* spp |  |
| 5 | Gynecology | *Citrobacter* spp, | *Staphylococcus* spp |
| 6 | Will chair | *E. coli* |  |
| 7 | Pre-term | *Edwardians* spp |  |
| 8 | Obstetric room | *Klebsiella* spp, *Acinitobacter* spp, | *Bacillus* spp *Enterococcus* spp  *Staphylococcus* spp |
| 9 | Delivery | *Klebsiella* spp, *Acinitobacter* spp, | *Bacillus* spp |
| 10 | Post-natal | *E. coli* | *Enterococcus* spp,  *Staphylococcus* spp |
| 11 | Male surgical | *E. coli , Klebsiella* spp, Non- lactose fermenter | *Staphylococcus* spp |
| 12 | Isolation (pediatric) | Non- lactose fermenter, *Enterobacter* spp | *Bacillus* spp, *Staphylococcus* spp |
| 13 | Pediatric ward | *Klebsiella* spp, Non- lactose fermenter, *Enterobacter* spp |  |
| 14 | stabilization phase I | *Klebsiella ozene*, 2Non- lactose fermenter | *Bacillus* spp  *Staphylococcus* spp |
| 15 | toilet door | *E. coli* |  |
| 16 | Pediatric OPD | 2 Non- lactose fermenter | *Bacillus* spp  *Staphylococcus* spp |
| 17 | Adult OPD | Non- lactose fermenter, *Klebsiella* spp | *Staphylococcus* spp |
| 18 | Reception | Non- lactose fermenter | *Staphylococcus* spp |
| 19 | Laboratory | *E. coli* | *Staphylococcus* spp |
| 20 | Female surgical ward | *Klebsiella* spp, *Enterobacter* spp, | *Staphylococcus* spp |
| 21 | Wards Door hands | *Klebsiella* spp, | *Staphylococcus* spp |
| 22 | Psychiatric room | *Enterobacter* spp | *Staphylococcus* spp |
| 23 | Female medical ward | *Enterobacter* spp | *Staphylococcus* spp |
| 24 | Staff cafeteria | *Enterobacter* spp | *Staphylococcus* spp |
| 25 | Pharmacy | Non- lactose fermenter |  |
| 26 | Autoclave |  |  |
| 27 | HC Adult OPD | Non- lactose fermenter | *Staphylococcus* spp |
| 28 | HC Emergency OPD | *E. coli*, Non- lactose fermenter |  |
| 29 | HC laboratory | *Acinitobacter* spp | *Staphylococcus* spp |
| 30 | HC ART clinic | *E. coli*, Non- lactose fermenter |  |
| 31 | HC EPI | Non- lactose fermenter | *Staphylococcus* spp |
|  | **Water samples** |  |  |
| 32 | Bidre hospital Borehole (Water) | Non- lactose fermenter, |  |
| 33 | Bidre health center water from tanker | Non- lactose fermenter, *Citrobacter* spp |  |

ART, antiretroviral therapy; EPI, expanded programme on immunization; HC, health center; ICU, intensive care unit; OPD, outpatient department

**Figure S1** Antibiotic susceptibility profile and multidrug resistance patterns in DoyoGena (A), Bulle (B), and Bidre (C)

**B**

**C**

**A**

|  |  |
| --- | --- |
| AMP | 67.7 |
| AMC | 27.3 |
| PTZ | 2.2 |
| CZO | 53.3 |
| CXT | 46.7 |
| CAZ | 28.9 |
| CEF | 24.4 |
| CTX | 46.7 |
| CIP | 21.2 |
| NA | 13.3 |
| CHL | 13 |
| COT | 52.8 |
| AMK | 10.9 |
| MER | 0 |
| TTC | 30.1 |
| PEN | 62.5 |
| OXA | 50 |
| MDR | 42.9 |

|  |  |
| --- | --- |
| AMP | 63.6 |
| AMC | 68.9 |
| PTZ | 28.9 |
| CZO | 86.6 |
| CXT | 57.8 |
| CAZ | 44.4 |
| CEF | 44.4 |
| CTX | 51.1 |
| CIP | 22.8 |
| NA | 26.7 |
| CHL | 36.8 |
| COT | 70.2 |
| AMK | 2.2 |
| MER | 0 |
| TTC | 54.4 |
| PEN | 100 |
| OXA | 83.3 |
| MDR | 71.1 |

|  |  |
| --- | --- |
| AMP | 57.1 |
| AMC | 44.9 |
| PTZ | 10.4 |
| CZO | 65.3 |
| CXT | 49 |
| CAZ | 34.7 |
| CEF | 30.6 |
| CTX | 44.9 |
| CIP | 20 |
| NA | 18.4 |
| CHL | 28.6 |
| COT | 54.4 |
| AMK | 14.3 |
| MER | 0 |
| TTC | 32.4 |
| PEN | 59.1 |
| OXA | 50 |
| MDR | 67.4 |

Percentage of resistance of 70 bacterial isolates identified from healthcare surface environmental and water samples of Bulle district of Gedo zone SPNN region of Ethiopia according to the CLSI disk diffusion breakpoints. Resistance was defined as isolates with intermediate resistance and complete resistance inhibition zone size. Antibiotics tested were ampicillin (AMP), amoxicillin-clavulanate (AMC), pepracillin with tazobactum (PTZ), cefazolin (CZO), cefuroxime (CXT), ceftazidime (CAZ), cefepime (CEF), cefotaxime (CTX), ciprofloxacin (CIP),naldixix acid (NA, chloramphenicol (CHL), Cotrimoxaxol (COT) Amikacin (AMK), Meropenem (MER),tetracycline (TTC), Penicillin (PEN) and oxacillin (OXA). MDR is to indicate the rate of Multidrug resistant bacterial isolates
